# Concurrent validity and between-device reliability of the Catapult Vector S8 GNSS device

**DOI:** 10.1101/2025.05.19.25327964

**Authors:** Susanne Ellens, Codey Moran, Matthew C. Varley

**Author notes:** **Corresponding Author:** Susanne Ellens, La Trobe University, School of Allied Health, Human Services and Sport, Melbourne, Australia.

## Abstract

This study assessed the concurrent validity and between-device reliability of the Catapult Vector S8 GNSS device for measuring distance, speed, acceleration and banded distance metrics. Twelve male sub-elite team sport athletes completed a testing protocol consisting of linear sprints, change of direction drills and a modified small-sided game. Validity was evaluated against criterion reference systems, a VICON motion capture system for most trials and a Stalker ATS radar for 50 m sprints, evaluated using root mean square error (RMSE) and mean bias. Between-device reliability was assessed using interclass correlation coefficients (ICC) and typical error (TE%). The Vector S8 demonstrated good validity with minimal errors for instantaneous distance (RMSE: 0.03 ± 0.01 m), speed (RMSE: 0.14 ± 0.05 m⸱s^-1^) and acceleration (RMSE: 0.29 ± 0.14 m⸱s^-2^). No overall bias was detected for instantaneous distance and speed, and the bias for acceleration (-0.016) was minimal. Accumulated distance showed a small underestimation across trials (mean bias -1.42%), with consistently low RMSE values (0.26-3.06 m), indicating high measurement precision. The 50 m sprint results showed similar validity, with minimal RMSE for instantaneous speed (0.14 ± 0.15 m⸱s^-1^) and acceleration (0.22 ± 0.22 m⸱s^-2^). Between device reliability demonstrated excellent agreement across all measured variables (ICC ≥ 0.90) with good precision (TE as CV <5%). No significant systematic bias was observed between devices for any variable (p > 0.05). This is the first study to validate the Catapult Vector S8 GNSS device, demonstrating that it is valid and reliable for measuring distance, speed and acceleration during sport-specific movements.

## Introduction

Global Navigation Satellite Systems (GNSS) are currently one of the most used technologies to track athletes (Crang et al., 2024; Delves et al., 2021; Ellens et al., 2022). The primary purpose of a GNSS is to quantify the external load of an athlete during training and matches (Akenhead & Nassis, 2016; Hudson et al., 2024). External load is determined by collecting kinematic data such as distance covered, sprints, accelerations and distance covered at certain speeds (Hudson et al., 2024). Practitioners and researchers use this information to enhance performance, support recovery and reduce injury risks (Akenhead & Nassis, 2016; Hudson et al., 2024). To allow meaningful interpretations of the GNSS data, it is important that the GNSS devices are both valid and reliable, to allow the user to make well informed decisions.

The validity of a GNSS device is defined as the degree to which the device accurately measures what it is intended to measure (Malone et al., 2017). This is typically evaluated by comparing the GNSS data to a criterion measure (gold standard). It is important that each measure of a GNSS device i.e., distance, speed and acceleration, is assessed in a validation study. A crucial aspect of validity assessment is the selection of an appropriate criterion reference measure, where measuring tapes and timing gates are the most commonly used criterion reference measures (Luteberget & Gilgien, 2020). A limitation of measuring tapes is that they do not capture the exact path travelled, especially by change of direction movements, potentially leading to inaccurate results. Furthermore, timing gates only provide average speeds over predefined distances (i.e., between two gates). Radar technology provides an alternative for instantaneous linear speed measurement, and is considered a gold standard for determining peak speed (Beato et al., 2018; Varley et al., 2012). However, radar technology cannot be used during sport-specific movements and is restricted to linear movements only. A 3D motion analysis system is considered a gold standard criterion for measuring instantaneous dynamic sport-specific movements and can be used during drills representing game-like situations (Luteberget & Gilgien, 2020). The reliability of a GNSS device refers to the reproducibility of measures on repeat occasions (Hopkins, 2000). When measurements of numerous GNSS devices are compared, which for example happens when comparing data of a squad of players, between-device reliability is important (Beato et al., 2018; Crang et al., 2021a; Scott et al., 2016). Between-device reliability is defined as the consistency of measurements between different devices when measuring the same movement (O’Donoghue, 2012).

Several factors influence the validity and reliability of GNSS devices. The sampling frequency of the GNSS device is a crucial factor affecting the validity and reliability, where higher sampling frequencies (≥ 10 Hz) have reported to be more accurate compared to GNSS devices with a lower sampling frequency (Beato et al., 2018; Crang et al., 2021b). The number of satellites and the positioning of the satellites interacting with the GNSS devices influences accuracy (Scott et al., 2016). The quality of the positioning of the satellites connected to the GNSS devices is defined by the horizontal dilution of precision (HDOP) (Specht, 2022). A HDOP value of ≤ 1 represents the ideal positioning of satellites in the sky and results in greater accuracy, this also requires a low variability and high mean number (≥ 12) of satellites (Specht, 2022). Multi-GNSS devices can connect to multiple satellite constellations (e.g., global positioning system (GPS), Galileo, GLONASS) concurrently, and therefore increasing the number of satellites it can connect to. An increase in error of distance and speed measures have been linked to a decrease in connected satellites and increase in HDOP values (Gray et al., 2010; Specht, 2022; Witte & Wilson, 2004).

Furthermore, changes in speed can compromise validity of GNSS devices with a sampling frequency < 10 Hz, where low accuracy is shown for movements with a high change in speed and frequent changes of direction (Buchheit et al., 2014; Jennings et al., 2010; Varley et al., 2012; Vickery et al., 2014). These older GNSS devices with a sampling frequency < 10 Hz utilise one satellite constellation (GPS), which also adds to the cause of decreased accuracy of the devices (Specht, 2022). However, more recent GNSS devices with a sampling frequency ≥ 10 Hz and connectivity to multiple satellite constellations, are less heavily influenced and have smaller error values for distance based measures for high changes in speed or frequent changes of direction movements (Bastida Castillo et al., 2018; Beato et al., 2018; Crang et al., 2024; Linke et al., 2018). However, the error in speed measures increases as the speed increases, where acceleration follows a similar pattern (Linke et al., 2018). Research also suggested that speed measured at lower speeds and likely higher accelerations are less accurate and slightly overestimated (Crang et al., 2024). Large differences in accuracy are present between GNSS device brands and models (Beato et al., 2018; Crang et al., 2021b). Therefore, validity and reliability studies should be conducted when new GNSS devices or models are released.

The validity and reliability of the Catapult S8 GNSS device have not yet been examined in the literature. Catapult is one of the most widely used GNSS device brands in high performance sport and research (Crang et al., 2024; Delves et al., 2021; Ellens et al., 2022), making its validity and reliability important for practitioners and researchers. Therefore, the aims of this study were to (I) investigate the concurrent validity of distance, speed and acceleration data and (II) between-device reliability of the Catapult Vector S8 GNSS device. Validity was established by comparing the measures to a criterion reference system during small-sided games, linear and change of direction drills.

## Methods

### Participants

Twelve male sub-elite soccer and Australian football players (mean ± SD; age: 21.4 ± 2.6 years, height: 184.4 ± 7.5 cm, body mass: 83.7 ± 13.8 kg) volunteered to participate in this study. Participants were recruited for this study in the period from 23-Sep-2024 until 21-Oct-2024. Prior to participation, all participants were informed about the risks and benefits of the research, before signing informed consent. The participants wore their normal training clothes during data collection, which took place over two days (one day apart). On each single day, six players participated. All sessions took place after sunset using floodlights on an empty and open (not surrounded by stands or buildings) grass soccer field. The weather was dry and windless with temperatures of 14° ± 1° Celsius. All procedures of this study were approved by the Human Research Ethics Committee of La Trobe University (reference number: HEC24295).

### Validated system

Catapult Vector S8 10 Hz GNSS devices (Firmware: 0.3.0; Catapult Sports, Melbourne, Australia) were used for this study. The GNSS devices provided position (longitude and latitude), distance (m), speed (m·s⁻¹) and acceleration (m·s⁻^2^) data which were processed and downloaded using the manufacturer’s software (OpenField, version 3.13.0, Catapult Sports, Melbourne, Australia). The manufacturer’s software processed data were used for this study, as this is the data used by practitioners. Distance was determined based on positional differentiation and the speed was doppler derived speed, which refers to speed measured by the shift in satellite signal frequency resulting from the movement of the GNSS device (Schutz & Herren, 2000). The acceleration data is subsequently derived from the doppler derived speed. Eleven different GNSS devices were used for this study. The GNSS devices were positioned in the manufacturer’s vest and placed between the participants shoulder blades. All GNSS devices were turned on 15 min prior to data collection to ensure each unit had a satellite lock. The study sample had an average ± SD horizontal dilution of precision (HDOP) of 0.99 ± 0.06 and 57.3 ± 1.78 number of satellites.

### Reference systems

A 3D infrared camera-based motion analysis system (VICON, Oxford, UK) capturing data at 100 Hz was used to determine the criterion reference for distance (m), speed (m·s⁻¹) and acceleration (m·s⁻^2^) data. The setup consisted of twenty-two cameras (ten Vantage 16 cameras, twelve Vantage 5 cameras, Software: Nexus 2.16) which were evenly spread around a 10 x 20 m capture area (Fig 1). Reflective markers with a diameter of 40.0 mm were used to ensure stable marker recognition within the capture area (10 x 20 m). Each participant in a session had a unique markup consisting of five markers, as can be seen in Fig 2. Only the marker corresponding to the location of the GNSS device was used for analysis. All other markers contributed to the unique markup and assisted in differentiating participants when multiple individuals were present in the capture area at the same time. Each marker was positioned on the participants skin using double sided tape and adhesive surgical tape on one of the following locations: middle of the GNSS device on the outside of the manufacturers vest (GNSS), right acromion (RACR), left acromion (LACR), right anterior superior iliac spine (RASI), left anterior superior iliac spine (LASI), right posterior superior iliac spine (RPSI), left posterior superior iliac spine (LPSI), sternum jugular notch (STJN), thoracic spine vertebra 12 (T12), lumbar spine vertebra 4 (L4), 1 cm below/inferior to the navel (NAVEL). The raw motion analysis data were filtered using a zero-lag fourth order low pass Butterworth filter with a 3Hz cut-off frequency which was determined based on residual analysis and choosing an equal balance between signal distortion and the amount of noise allowed through (Linke et al., 2018; Winter, 2009). Gaps in the data ≤ 5 samples (0.05 s) were filled using spline interpolation. Gaps that were ≥ 5 samples were excluded from analysis. The X and Y coordinate data corresponding to the middle of the GNSS device were exported and used for analysis. The z-coordinates (vertical axis) were excluded from analysis, as the S8 GNSS device does not collect data in the vertical axis.

**Fig 1.**
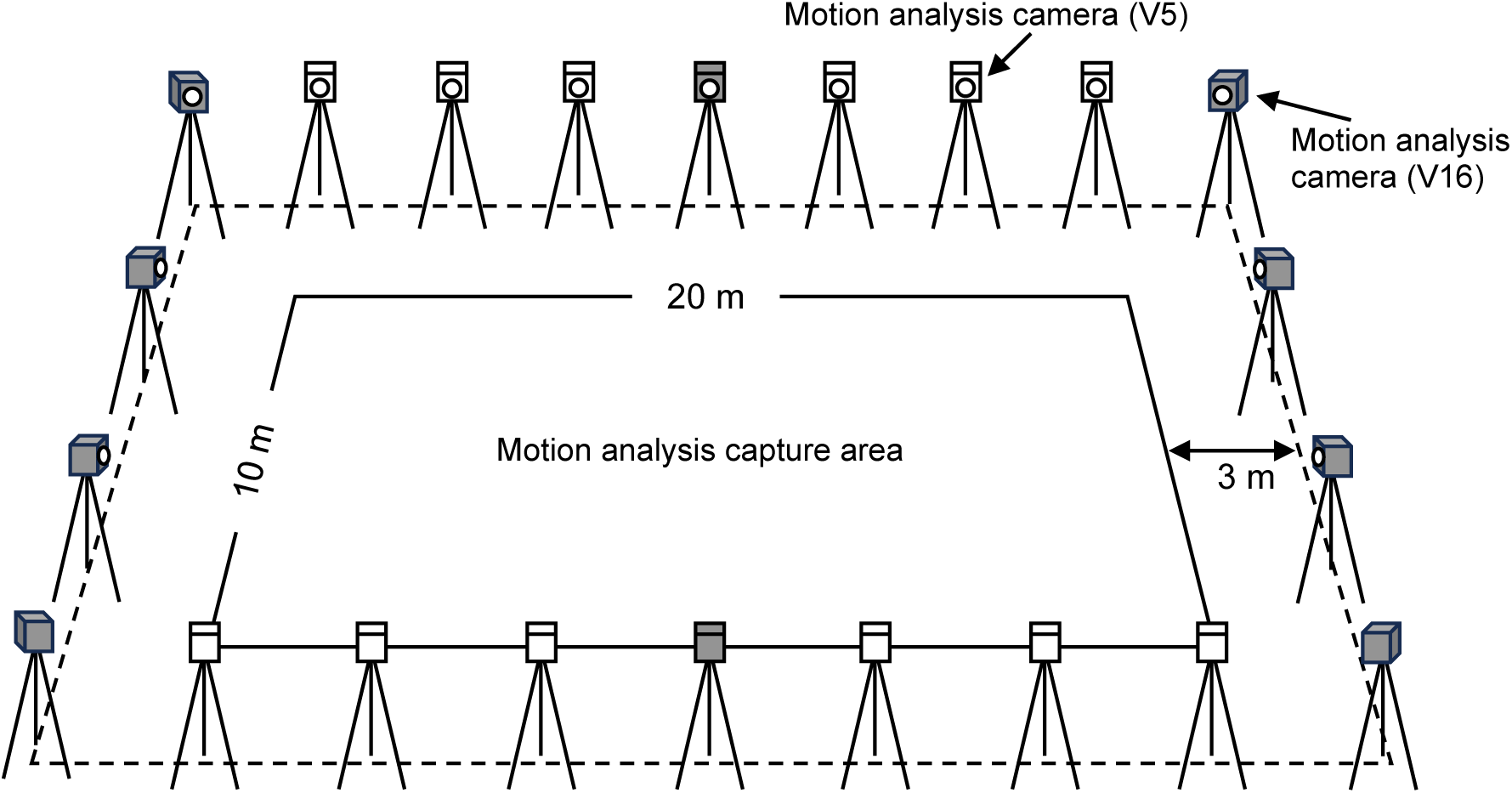
Motion analysis camera setup. Motion analysis camera setup around the capture area where all trials were performed. The grey-filled motion analysis cameras refer to the Vantage 16 (V16) model, while the white-filled cameras represent the Vantage 5 (V5) model.

**Fig 2.**
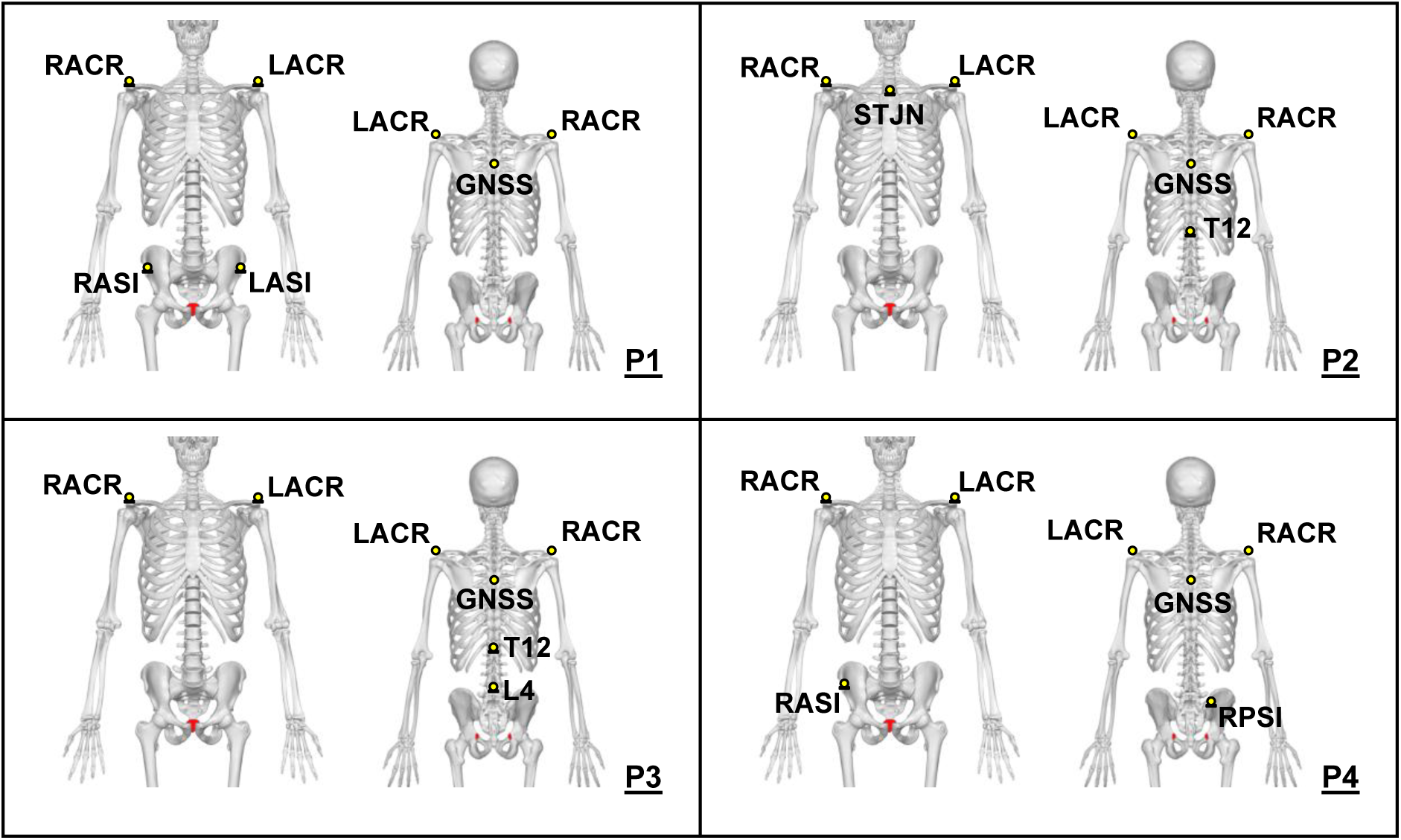
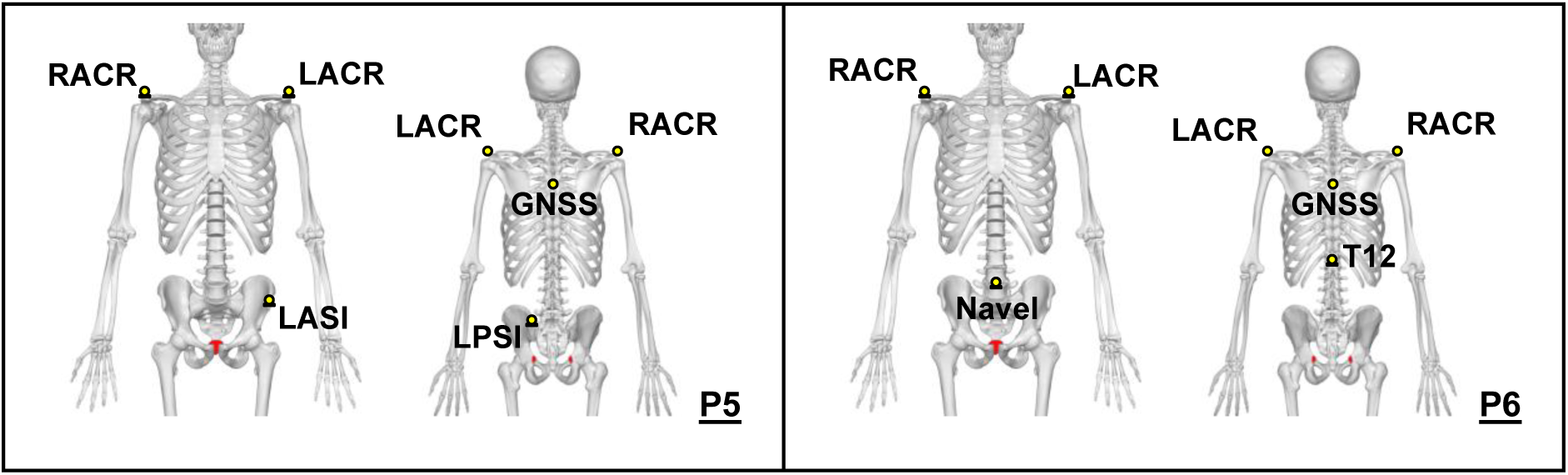
Participant markup. Unique markup and positions of the reflective markers for each of the six participants (P1-P6) in a data collection session.

Criterion measure of maximal speed, captured over 50 meters, was measured using a Doppler radar (Stalker ATS II, Plano, TX, USA, 47Hz), as the motion capture area was not large enough to permit a 50 m sprint. Raw, unprocessed radar speed data were exported via proprietary software (Stalker ATS 5.1.1). The radar has a reported ± 0.045 m⸱s^-1^ accuracy within a field of 5 degrees from the radar (Applied Concepts, 2018).

### Movement trials

#### Validity

Seven different movement trials, illustrated in Fig 3, were performed by the participants with 60 seconds of passive recovery between trials. The following movement trials were included in the data collection: Change of direction trials of 45°, 90° and 180° consisted of a 5 m sprint into the change of direction into 5 m sprint; Linear runs of 10 m, 20 m and 50 m; Circuit with a flying start consisting of sharp turns, accelerations and decelerations, totalling 70 m; Modified small sided game (SSG) of 2 min, 3vs3 where it was the objective to pass a tennis ball among teammates and score points. Players needed to complete at least five successful passes before attempting to score by catching the ball in a 1×1 m goal area at their opponent’s end. Running with the ball was not allowed, forcing the players to create space by accelerating, decelerating and performing changes of direction at a high intensity. It was instructed and encouraged during the SSGs that players without the ball keep moving at a high intensity. All participants started and ended each trial in a stationary position and were instructed to perform to their maximal ability, with each trial performed twice.

**Fig 3.**
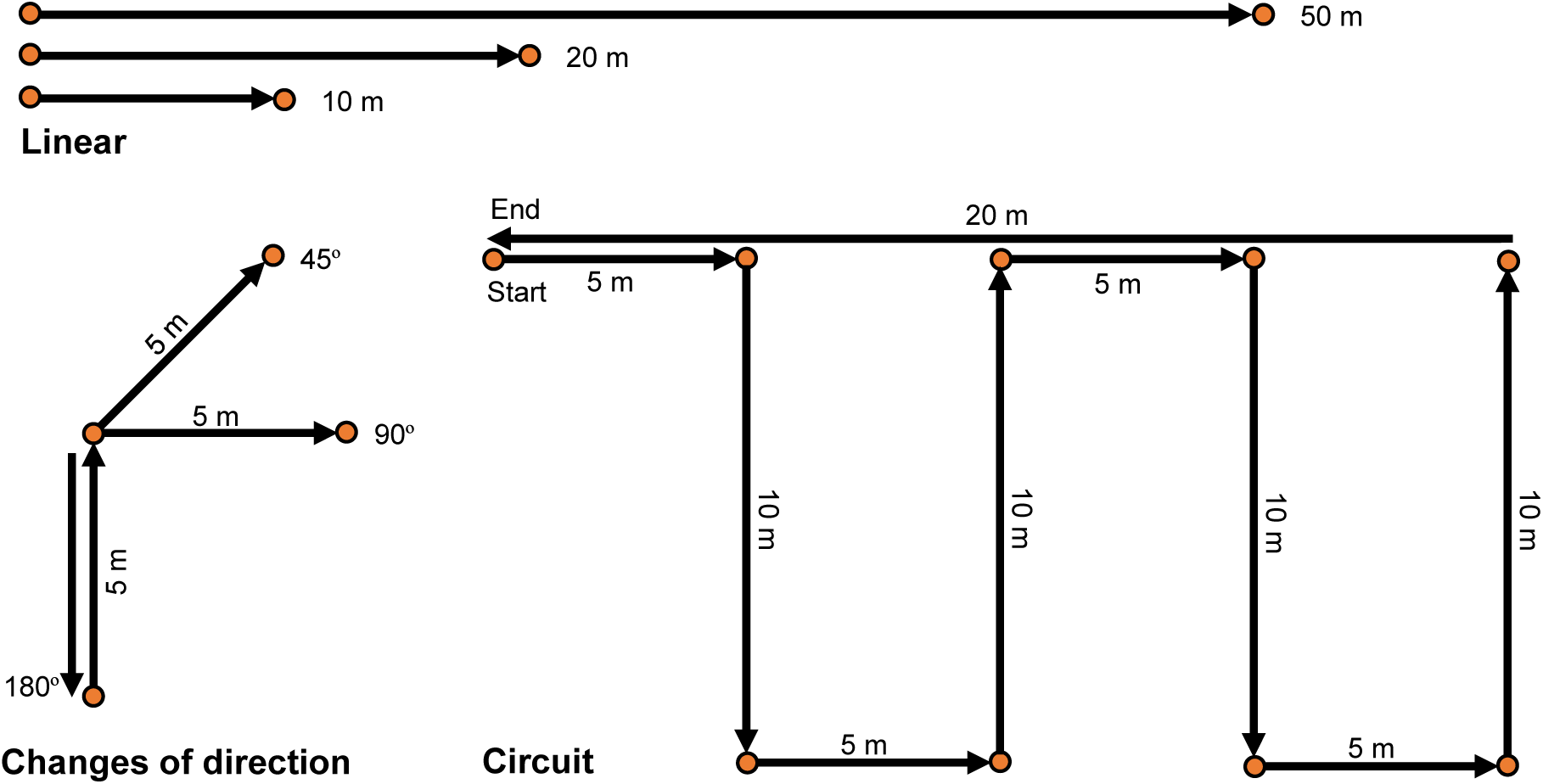
Illustration of movement trials. Linear, changes of direction and circuit exercises performed during the validation testing.

The total amount of exercises included for analysis consisted of 24 COD 45°, 24 COD 90°, 23 COD 180°, 24 linear 10 m, 24 linear 20 m, 17 linear 50 m, 18 circuit, 16 SSG. A total of 21, 50 m sprints were conducted, three participants performed the 50 m once. Four 50 m trials were excluded due to radar device measurement errors. For the motion analysis data, 14 data files (6 circuit and 8 SSG) contained data gaps and were excluded from analysis.

#### Between-device reliability

Participants completed a standardised intermittent running protocol of ∼139m while wearing two GNSS devices positioned between the scapulae in a custom multi-pouch garment. Each participant performed three trials of the protocol, with 60 seconds of passive recovery between trials, resulting in 36 trials included for analysis. The protocol included high-speed running (> 5 m⸱s^-1^), maximal efforts, jogging, walking, and changes of direction, as shown in Fig 4.

**Fig 4.**
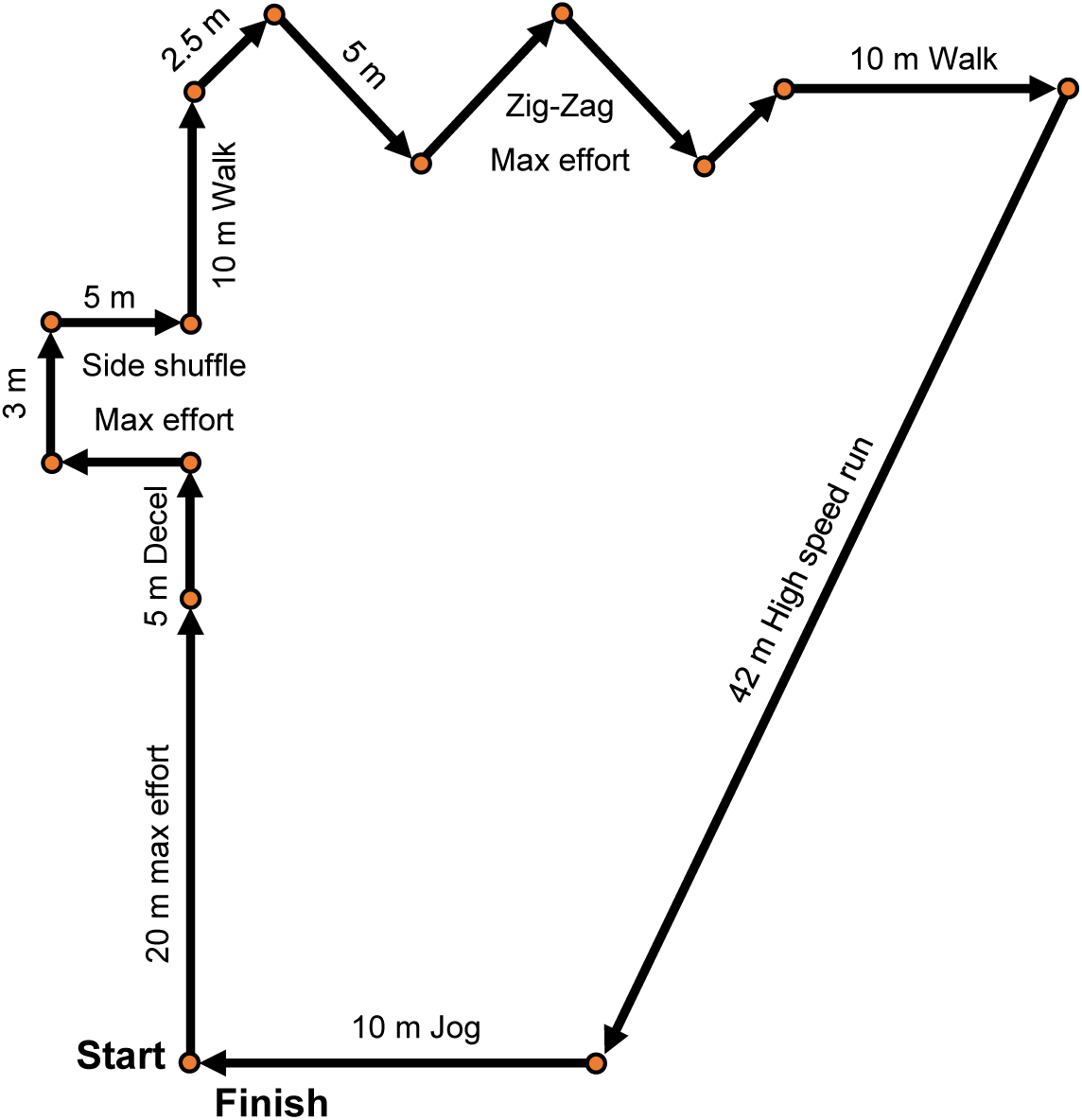
Between-device reliability protocol. Standardised intermittent running protocol for the between-device reliability trials. Decel = deceleration, High speed run > 5 m⸱s^-1^.

### Data analysis

The speed of the motion analysis data was calculated by differencing the positional data and applying the same filter as used in the manufacturer software (OpenField, version 3.13.0) on the GNSS data. Previous research has shown that different filters applied to the GNSS data can heavily influence and affect data (Delaney et al., 2019; Delves et al., 2022; Ellens et al., 2024; Thornton et al., 2019; Varley et al., 2017). It was therefore chosen to use the same filtering practises on the criterion data as those used on the GNSS data. This ensured that any differences present in the data were not due to the filtering practices used, but due to differences in the collected raw data. The filtering information was provided to the researchers by the manufacturer; however, details are not included here due to the manufacturer intellectual property. Similarly, acceleration was calculated by differencing the speed data and filtered using the manufacturer’s specifications. The derived data were then synchronised to the GNSS data by cross correlating the speed signals to find the time offset that maximised the correlation (Buck et al., 2002). The data were then further synchronised by shifting the speed signal forwards and backwards by 50 data points intervals in one step increments. The root mean squared error (RMSE) was established at each shift, where the shift with the lowest RMSE was used to synchronise the data. The synchronised motion analysis data were down sampled to 10 Hz to match the GNSS data and were used for analysis.

Raw, unprocessed radar speed was filtered using the same method as the manufacturer software. Acceleration was derived by differentiating the filtered speed, which was also processed based on the manufacturer’s specifications. The filtered radar data were down sampled to 10 Hz and synchronized with the Catapult data by cross correlating the speed signals to determine the time offset that maximized correlation. All data processing and analysis were conducted using the R statistical programming language (version 4.2.0) and the gsignal package.

### Statistical analysis

#### Validity

The magnitude of the error and the relationship between the GNSS data and the criterion for distance, speed and acceleration were assessed using root mean square error (RMSE), mean bias and mean bias percentage error (%) (Scott et al., 2016). The mean bias percentage error is defined as percentage difference in relation to the criterion reference measure, calculated for each trial and averaged across each respective movement trial category. To quantify the difference between the criterion and GNSS for instantaneous distance, speed and acceleration, RMSE and mean bias were calculated for each trial by evaluating the instantaneous error (GNSS – Criterion) at each data point across the synchronized 10 Hz time series and presented as RMSE (mean ± standard deviation) and Mean Bias ± 90% confidence interval (CI).

#### Between-device reliability

The between-device reliability of the Vector S8 device for measuring distance, speed and acceleration was assessed using the following metrics: Total distance (m), maximal speed (m⸱s^-1^), banded distance (m) for low-speed running (LSR = 0-5 m⸱s^-1^) and high speed running (HSR > 5 m⸱s^-1^), maximal acceleration (m⸱s^-2^), maximal deceleration (m⸱s^-2^) and acceleration load defined as the accumulated absolute acceleration values. The reliability was evaluated using typical error (TE) expressed as a coefficient of variation (CV, 90% CI) and intraclass correlation coefficients (ICCs, 90% CI) using two-way mixed-effects model, absolute agreement, single measurement (Atkinson & Nevill, 1998). Reliability was classified as good if CVs were <5%. ICCs were interpreted as follows: poor (<0.5), moderate (0.5–0.75), good (0.75–0.9), and excellent (>0.9) (Portney & Watkins, 2015).

Systematic bias between devices was assessed using paired t-tests, with statistical significance set at p < 0.05. Normality was tested using the Shapiro-Wilk test. If the assumption of normality was violated, comparisons were made using the nonparametric Wilcoxon signed-rank test. Agreement and variability between devices were further examined using Bland-Altman analysis, which provided mean bias and 90% limits of agreement.

## Results

### Validity

The validity results of the GNSS devices compared to the motion analysis criterion for instantaneous data are presented in Table 1, and for accumulated distance data in Table 2. The overall RMSE for instantaneous distance (0.03 ± 0.01), speed (0.14 ± 0.05) and acceleration (0.29 ± 0.14) were minimal. There was no overall bias for instantaneous distance and speed, and the bias for acceleration (-0.016) was minimal. The difference between all instantaneous speed and acceleration measurements compared to the motion analysis criterion data are shown in Figs 5 and 6. A visual representation of the accuracy of speed data across movement trials with median mean difference and 95^th^ percentile mean difference are shown in Figs 7 and 8.

**Fig 5.**
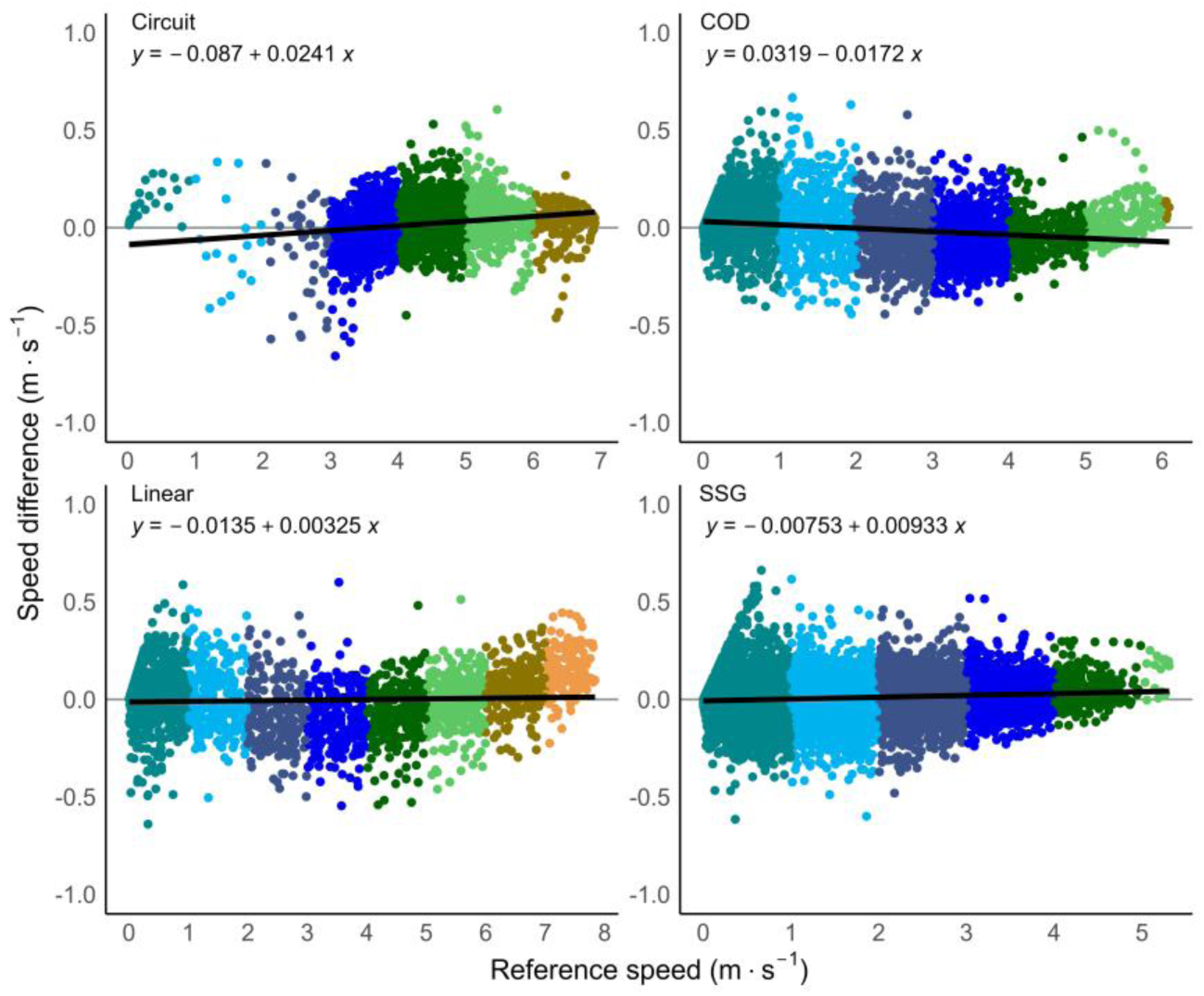
Difference in speed by movement category. Instantaneous criterion reference speed (m⸱s^-1^) of the motion analysis (MA) data compared to the global navigation satellite system (GNSS) data, divided into speed thresholds and split by movement category. Speed difference = MA – GNSS data. COD = change of direction trials, SSG = modified small sided game trials.

**Fig 6.**
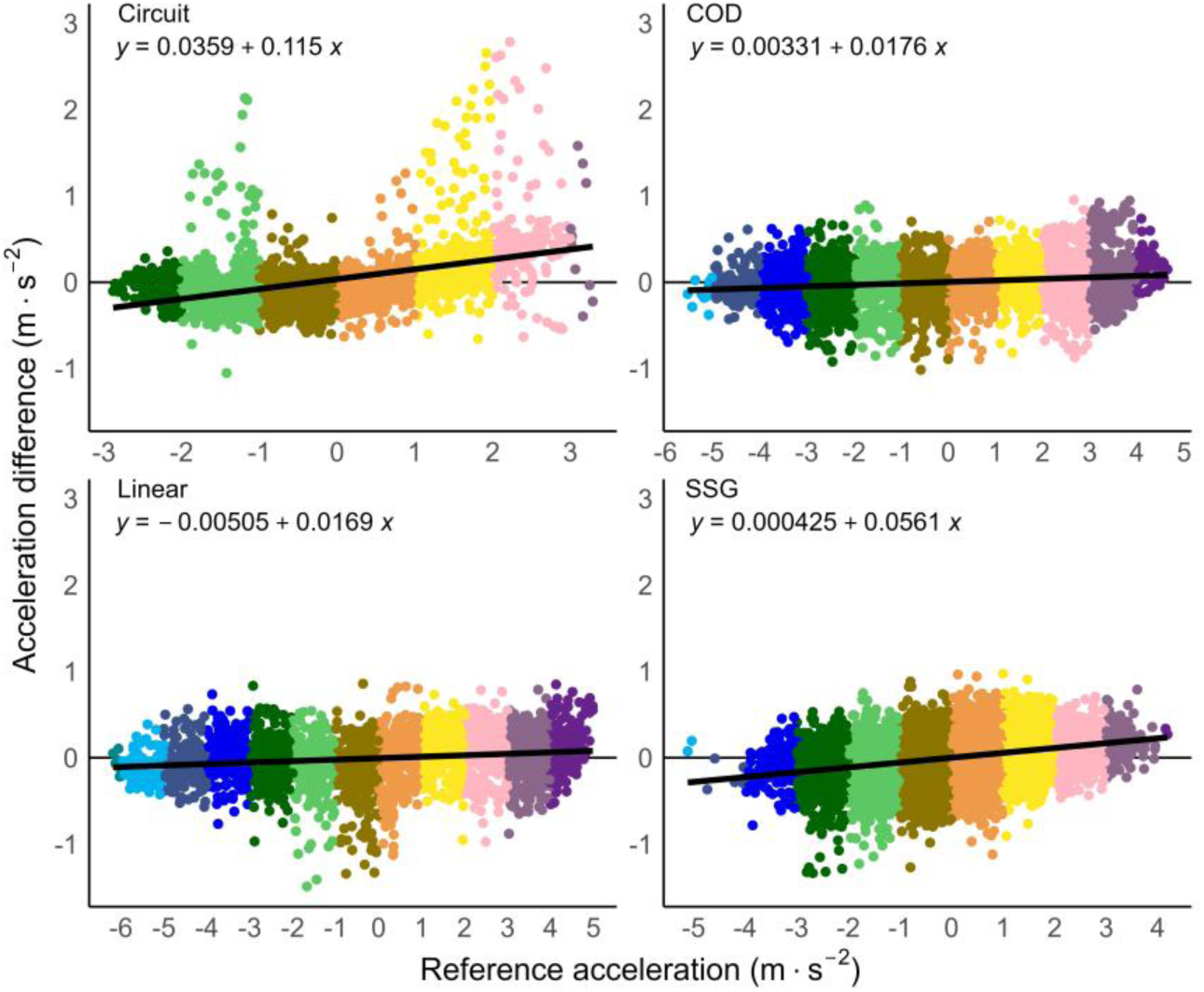
Difference in acceleration by movement category. Instantaneous criterion reference acceleration (m⸱s^-2^) of the motion analysis (MA) data compared to the global navigation satellite system (GNSS) data, divided into acceleration thresholds and split by movement category. Acceleration difference = MA – GNSS data. COD = change of direction trials, SSG = modified small sided game trials.

**Fig 7.**
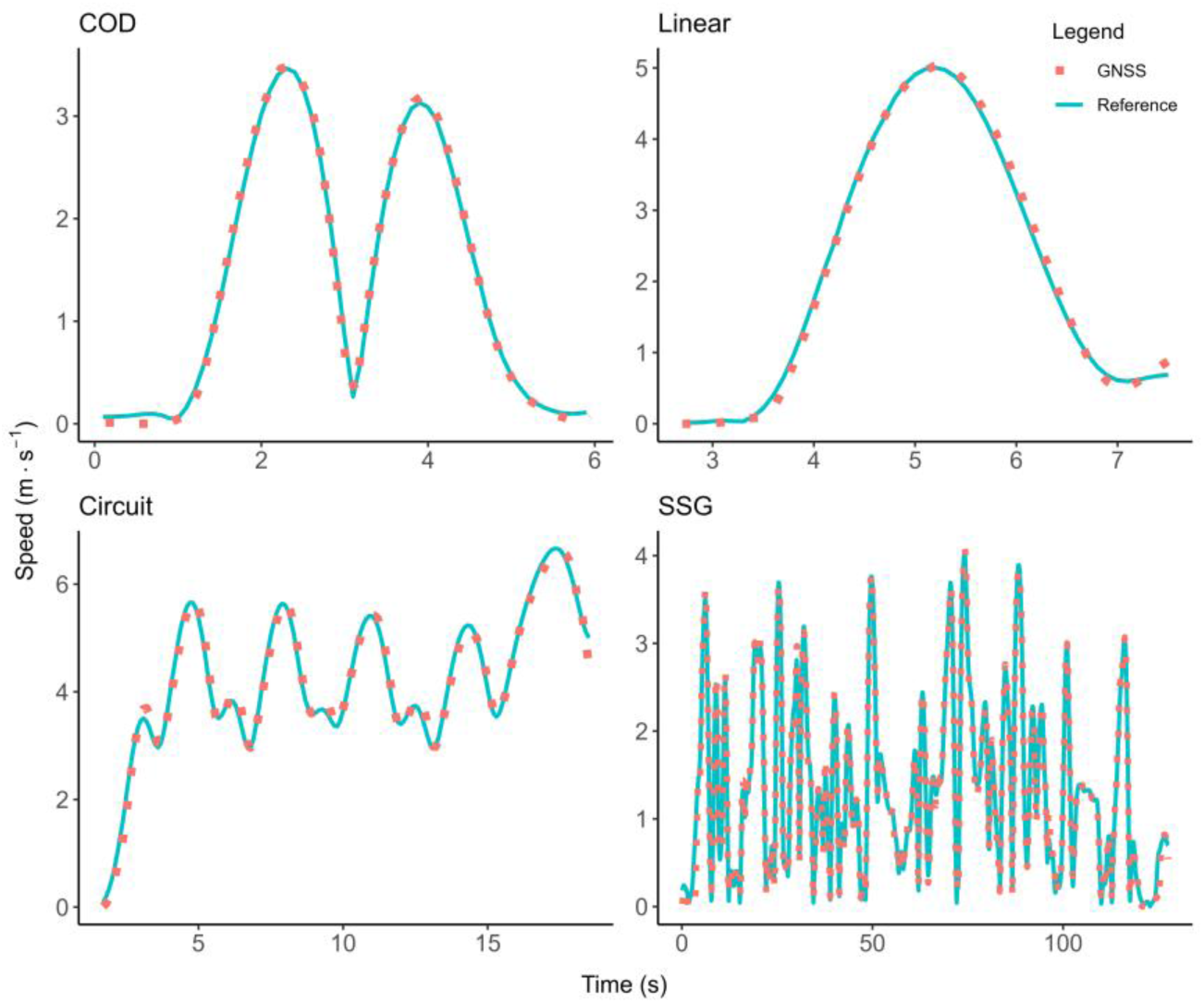
Median mean difference error speed traces. Accuracy of speed data across movement trials with a median mean difference error. The speed traces of the global navigation satellite data (GNSS – red dotted line) are plotted on top of the motion analysis reference speed data (blue line) to show alignment of the data.

**Fig 8.**
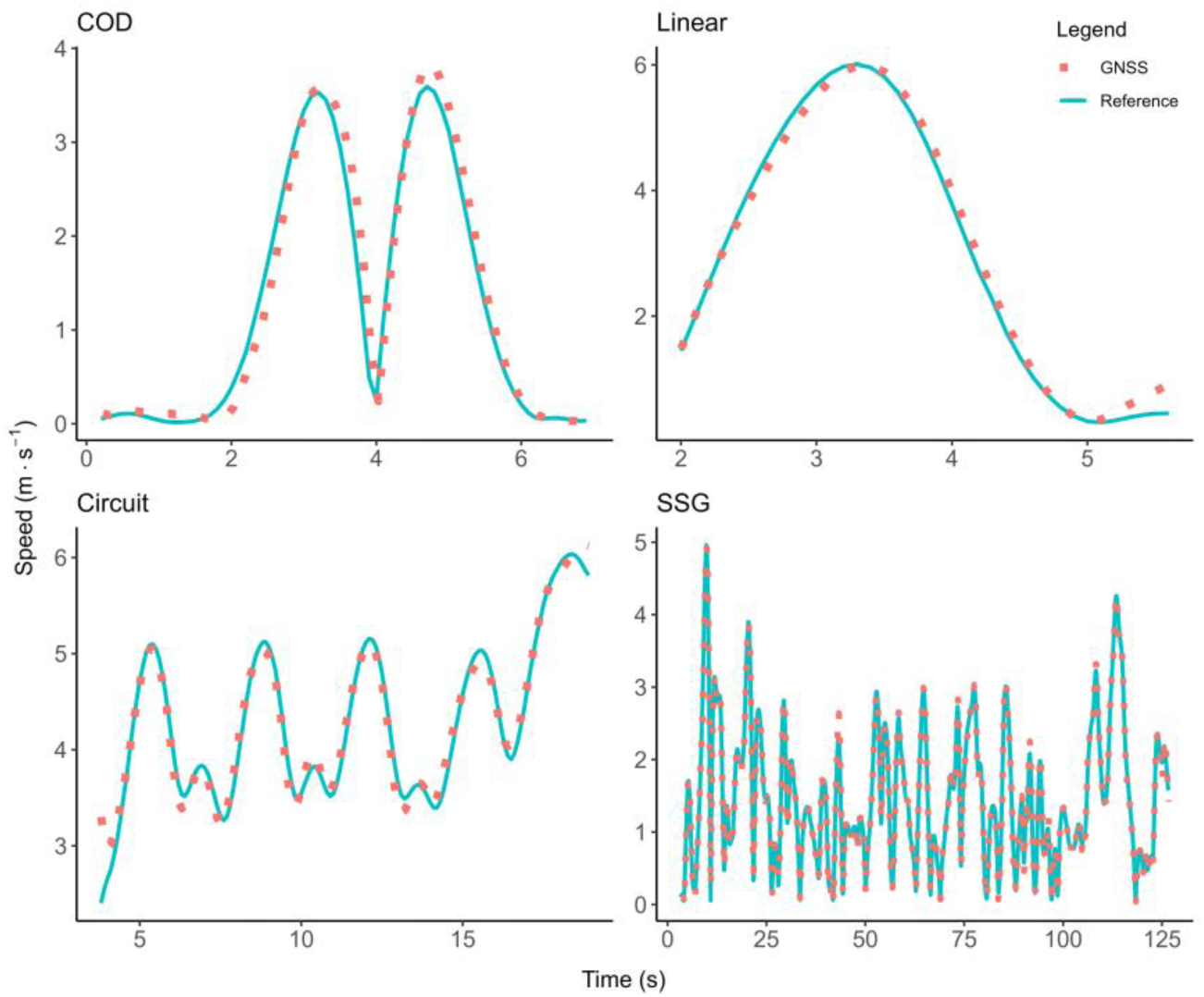
The 95^th^ percentile mean difference error speed traces. Visual representation of the accuracy of speed data across movement trials with a mean difference error in the 95^th^ percentile. The speed traces of the global navigation satellite data (GNSS, red dotted line) are plotted on top of the motion analysis reference speed data (blue line) to show alignment of the data.

**Table 1.**
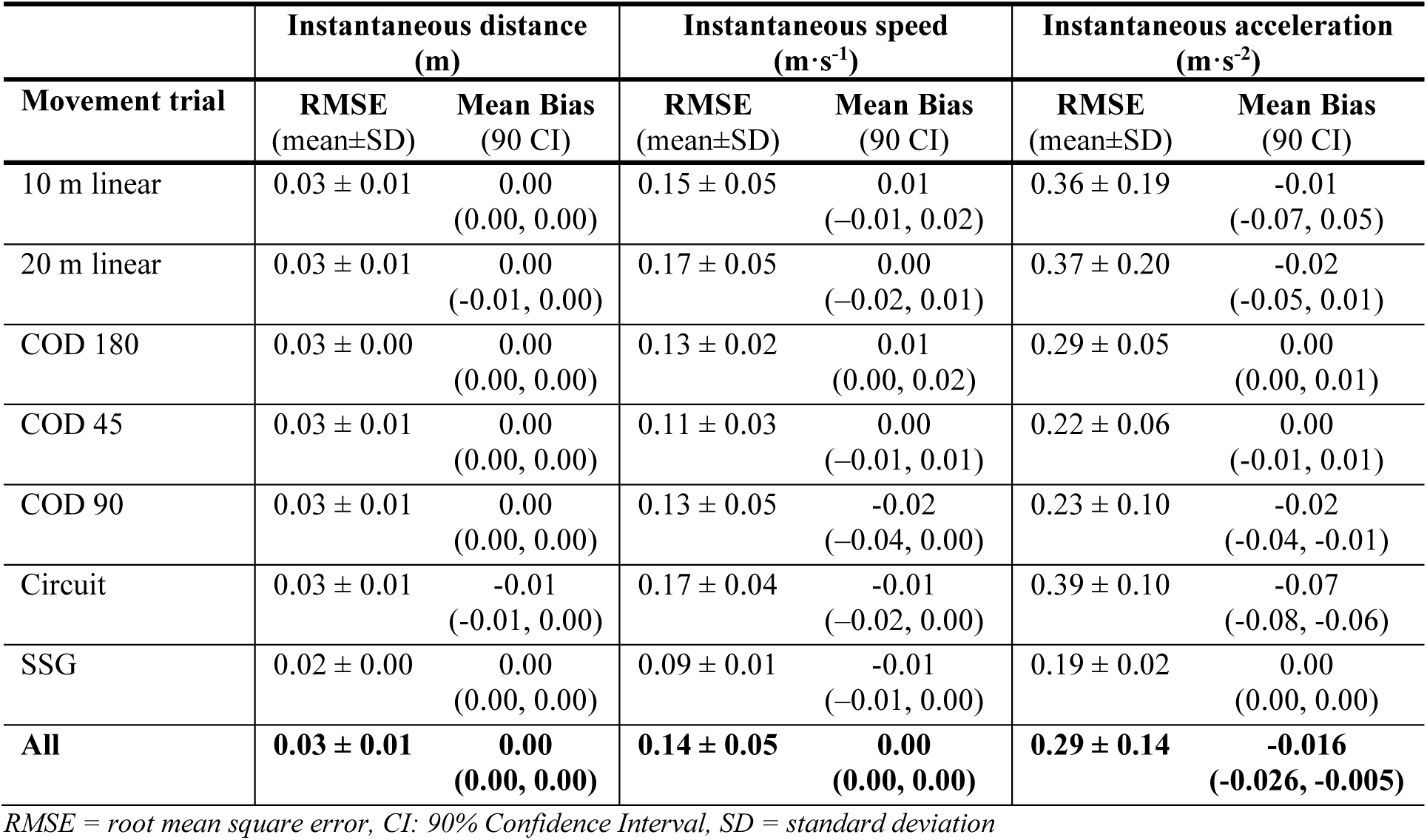
Validity results of instantaneous distance (m), speed (m⸱s^-1^) and acceleration (m⸱s^-2^) split by movement trial.

**Table 2.** Validity results of accumulated distance (m) split by movement trial.

| Accumulated distance (m) |  |  |  |  |  |
| --- | --- | --- | --- | --- | --- |
| Movement trial | GNSS<br>(mean ± SD) | Criterion<br>(mean ± SD) | RMSE | Mean Bias (m) | Mean Bias % |
| Linear 10m | 10.54 ± 0.48 | 10.69 ± 0.58 | 0.26 | -0.15 | -1.37 |
| Linear 20m | 20.13 ± 0.53 | 20.36 ± 0.60 | 0.33 | -0.23 | -1.16 |
| COD 180 | 9.29 ± 0.45 | 9.48 ± 0.48 | 0.29 | -0.19 | -2.09 |
| COD 45 | 12.18 ± 0.38 | 12.37 ± 0.36 | 0.26 | -0.18 | -1.52 |
| COD 90 | 10.12 ± 0.57 | 10.23 ± 0.49 | 0.30 | -0.11 | -1.18 |
| Circuit | 69.28 ± 4.06 | 70.12 ± 3.98 | 0.94 | -0.84 | -1.22 |
| SSG | 190.66 ± 12.92 | 193.39 ± 13.49 | 3.06 | -2.73 | -1.42 |

Criterion = 3D motion analysis criterion reference measure, global navigation satellite system (GNSS), RMSE = root mean square error.

The validity results of the GNSS devices compared to the radar for the 50 m linear run are presented in Table 3. The 50 m linear run had comparable results to the motion analysis trials, with minimal RMSE for instantaneous speed (0.14 ± 0.15) and acceleration (0.22 ± 0.22). The linear trials together with the circuit displayed had larger RMSE values on average compared to the change of direction trials, although not significantly different. The difference between all instantaneous speed and acceleration measurements compared to the radar criterion data during the 50 m linear trials are shown in Fig 9.

**Fig 9.**
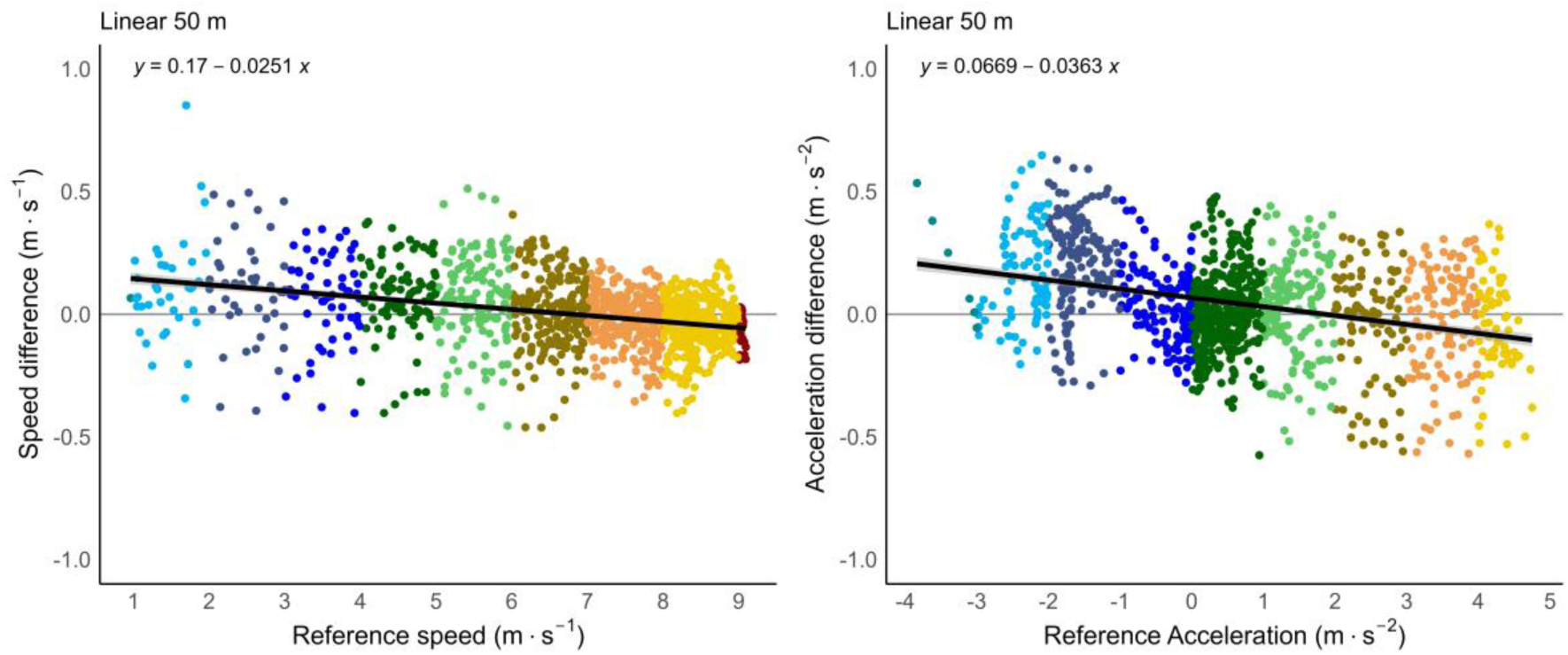
Difference in speed and acceleration of 50 m sprint trials. Instantaneous speed (left) and acceleration (right) error (Criterion – GNSS) of the GNSS during the maximal 50 m sprint trials.

**Table 3.** Validity results of the 50 m linear sprint trials.

| 50 m linear sprint trials |  |  |  |  |  |
| --- | --- | --- | --- | --- | --- |
|  | GNSS<br>(mean±SD) | Criterion<br>(mean±SD) | RMSE | Mean Bias | Mean Bias % |
| Peak speed ($\text{m} \cdot \text{s}^{-1}$) | $8.52 \pm 0.44$ | $8.54 \pm 0.44$ | 0.05 | -0.03 | -0.4% |
| Peak acceleration ($\text{m} \cdot \text{s}^{-2}$) | $4.23 \pm 0.26$ | $4.25 \pm 0.26$ | 0.17 | -0.02 | -0.5% |
| Instantaneous speed ($\text{m} \cdot \text{s}^{-1}$) | $6.72 \pm 1.88$ | $6.72 \pm 1.92$ | $0.14 \pm 0.15$ | 0.00<br>(-0.01, 0.01) | |
| Instantaneous acceleration ($\text{m} \cdot \text{s}^{-2}$) | $0.69 \pm 1.76$ | $0.65 \pm 1.82$ | $0.21 \pm 0.20$ | 0.04<br>(0.03, 0.05) | |

Criterion = Radar criterion reference measure, global navigation satellite system (GNSS), RMSE = root mean square error, RMSE is presented as the mean ± standard deviation, Mean Bias is presented with 90% confidence interval.

#### Between device reliability

Between device reliability results are presented in Table 4 and demonstrated excellent agreement across all measured variables (ICC ≥ 0.9) with good precision (TE as CV <5%). No significant systematic bias was detected between devices for any variable (p > 0.05).

**Table 4.** Between device reliability results.

|  | GNSS<br>device #1<br>(mean±SD) | GNSS<br>device #2<br>(mean±SD) | Mean Δ<br>(90% CI LOA) | p | ICC<br>(90% CI) | TE as CV%<br>(90% CI) |
| --- | --- | --- | --- | --- | --- | --- |
| Total distance (m) | 138.9 ± 5.2 | 139.0 ± 5.2 | 0.08<br>(-1.06, 1.22) | 0.48 | 0.99<br>(0.99, 1.00) | 0.34<br>(0.27, 0.41) |
| Max speed (m·s<sup>-1</sup>) | 6.94 ± 0.61 | 6.95 ± 0.61 | 0.01<br>(-0.08, 0.10) | 0.30 | 1.00<br>(0.99, 1.00) | 0.52<br>(0.42, 0.62) |
| LSR (m) | 87.9 ± 9.6 | 87.8 ± 9.3 | -0.09<br>(-1.85, 1.67) | 0.61 | 0.99<br>(0.99, 1.00) | 0.83<br>(0.67, 0.99) |
| HSR (m) | 50.9 ± 6.7 | 51.1 ± 6.3 | 0.16<br>(-1.49, 1.81) | 0.34 | 0.99<br>(0.98, 0.99) | 1.35<br>(1.09, 1.61) |
| Max acceleration (m·s<sup>-2</sup>) | 3.70 ± 0.38 | 3.67 ± 0.39 | -0.03<br>(-0.23, 0.18) | 0.23 | 0.95<br>(0.91, 0.97) | 2.39<br>(1.92, 2.85) |
| Max deceleration (m·s<sup>-2</sup>) | -3.27 ± 0.38 | -3.29 ± 0.38 | -0.02<br>(-0.19, 0.15) | 0.07 | 0.99<br>(0.98, 0.99) | -2.22<br>(-1.79, -2.66) |
| AccelerationLoad | 37.0 ± 2.9 | 37.1 ± 2.9 | 0.11<br>(-1.48, 1.70) | 0.48 | 0.95<br>(0.91, 0.97) | 1.78<br>(1.44, 2.13) |

\* $p < 0.05$ . Paired T-Test; ICC = interclass correlation coefficient, TE = Typical Error, CV = coefficient of variation, CI = 90 % confidence interval; global navigation satellite system (GNSS), LSR = 0-5 m·s<sup>-1</sup>, HSR = >5 m·s<sup>-1</sup>

## Discussion

The main findings of this study were that the Catapult Vector S8 GNSS device is valid for measuring instantaneous distance, speed, acceleration and peak speed and acceleration. Furthermore, the measures of total distance, maximal speed, low speed running (0-5 m⸱s^-1^), high speed running (> 5 m⸱s^-1^), maximal acceleration, maximal deceleration and acceleration load displayed excellent reliability between GNSS devices.

The validity of the Catapult Vector S8 10 Hz GNSS device appeared suitable for measuring instantaneous speed. The validity results are comparable to previous research that used older 10 Hz Catapult devices (Crang et al., 2024). The current study had an average RMSE for instantaneous speed of 0.14 compared to 0.15 of the previous similar research. Previous research has shown that instantaneous speed is affected by high changes in speed which mainly occur at lower speeds (Akenhead et al., 2014; Crang et al., 2024; Varley et al., 2012). The change in speed from sample to sample can be largest during lower speeds as more force can be applied to the ground compared to higher speeds (Morin et al., 2011), impacting the validity of the speed data. This is similar to the results of the current study, where data involving lower speeds had reduced accuracy compared to the criterion reference (Fig 5 and Fig 9). However, the error in this study for these datapoints were low (mean bias instantaneous speed < 0.02 m⸱s^-1^) and when averaged across all time-series, the errors were negligible (mean bias instantaneous speed = 0.00 m⸱s^-1^) and are unlikely to affect derived metrics. For example, maximal speed values expected during a soccer game are ∼9 m⸱s^-1^ (Morin et al., 2021), a RMSE of 0.14 is only 1.5% away from the potential maximal speed value. Practitioners can be confident in the validity of the instantaneous speed data derived from the Catapult Vector S8 GNSS device, as error values were close to zero across all conditions.

The instantaneous acceleration of the GNSS across all movement trials has shown to offer suitable validity. Compared to previous research (Crang et al., 2024), the current study had an average RMSE for instantaneous acceleration of 0.29 compared to 0.39 of the previous similar research. However, the scatterplots in Fig 6 demonstrate a positive trend for instantaneous acceleration where the error increased with acceleration. Both high positive and high negative acceleration values tended to be overestimated, with the latter being underestimated in magnitude. This suggests that the GNSS devices overestimated instantaneous acceleration values during rapid changes of speed. These findings align with observations of previous studies (Akenhead et al., 2014; Crang et al., 2024; Varley et al., 2012) that highlighted high instantaneous acceleration values are overestimated and likely affected due to the larger point to point differences (which for example happens when starting to move from standstill, or performing an abrupt stop from high speed). It is important to note that the acceleration data is derived from the speed data, and the accuracy of the acceleration data is therefore directly related to the accuracy of the speed data. As differentiation amplifies noise, the error in acceleration data (RMSE 0.29 in this study) is also expected to be greater than the speed data (RMSE 0.14 in this study). The trend seen in the plots reinforces that errors in instantaneous acceleration data are linked to both the underlying speed data and the nature of the movement. Despite this, the overall mean error remains low, practitioners and researchers can therefore still confidently rely on instantaneous acceleration data and average acceleration metrics, which are valid and representative of performance across linear and change of direction movement sequences.

The measurement error between the radar and GNSS data when measuring maximal speed and acceleration was minimal. The accuracy of GNSS devices to measure maximal speed has been linked to the sample rate, where an increase in sample rate (5 to 20 Hz) was linked to decreasing typical errors (5.1 to 2.3%) (Nagahara et al., 2017). A more recent paper using a 25-Hz GNSS device showed improved validity with typical errors of 0.5% and a mean bias of -0.07 m⸱s^-1^ for maximal speed (Dennison et al., 2025). Furthermore, research using the previous Catapult S7 GNSS devices sampling at 10 Hz have shown to achieve comparable accuracy in maximal speed values to the 20-25 Hz devices, with reported mean bias for peak speed of -0.05 to -0.08 m⸱s^-1^ (Clavel et al., 2022; Crang et al., 2024). The Catapult S7 GNSS devices also showed good accuracy for maximal acceleration, with reported mean bias of -0.25 m⸱s^-2^ (Crang et al., 2024). The results of the current study showed further improvements in validity with a low mean bias for maximal speed of -0.03 m⸱s^-1^ and maximal acceleration of -0.02 m⸱s^-2^. The improved validity of the new Catapult S8 GNSS device compared to the previous Catapult S7 GNSS device for peak speed and acceleration could be due to the improved satellite connectivity of the S8 devices. The current study had an average of 57 connected satellites, compared to 15-16 satellites reported in similar previous research using the S7 device (Clavel et al., 2022; Crang et al., 2024). In addition, the reliability of the maximal speed and acceleration were excellent (ICC 0.95-1.0, see Table 4), therefore practitioners and researchers can be confident in their measured maximal speed and acceleration for each GNSS device and between GNSS devices.

The accuracy of the GNSS in measuring accumulated distance showed a small underestimation across all movement trials (mean bias -0.11 to -2.73 m; -1.16 to -2.09%). The RMSE values remained low across trials, ranging from 0.26 m for 10 m linear trials to 3.06 m in the modified SSG, indicating high measurement precision. The modified SSG, despite being the most complex and dynamic trial, still had a low RMSE of 3.06 m compared to similar research (Linke et al., 2018) who reported a RMSE of 4.90 m for a similar GNSS device and comparable total distance trial (193 m vs 224 m). Furthermore, the RMSE increased for trials with longer (modified SSG) compared to shorter (COD 45°) distances (see Table 2), but the relative measurement error stayed consistent across trials (mean bias %). Thus, although the error increased as total distance increased (as seen in longer trials like the modified SSG), the degree to which the GNSS underestimated remained predictable and consistent. To put the error in context, a professional soccer player has an average total match distance of 11.4 ± 1.0 km (Di Salvo et al., 2007). An underestimation of -1.42% would correspond to a difference in total distance of 0.16 km, which is 0.16 standard deviations (1.0 km) and is considered a small and negligible difference (Cohen, 2013). Overall, the GNSS device provided improved accuracy for accumulated distance, where an average underestimation of -1.42% is expected. The underestimation of distance could be due to the ability of a motion analysis system to detect microscopic movements, which do not result in a noticeable change of position of the participant. These microscopic movements, which may occur as a result of subtle body sway, can only be measured using highly sensitive devices such as a motion analysis system (Linke et al., 2018). However, GNSS devices are not designed to measure athlete movements at this level of precision and the results of the current study might therefore be underestimated.

The between-device reliability showed excellent agreement across all measured variables (ICC ≥ 0.9) with good precision (TE as CV <5%) see Table 4. The results also showed greater between-device reliability than previous research for measures of total distance, maximal speed, maximal acceleration and deceleration, where the current study had ICC values of 0.95-1.0 compared to 0.2-0.99 in the previous research (Clavel et al., 2022; Jackson et al., 2018). A custom vest was used for the reliability data collection of the current study, which could result in better reliability results. The custom vest ensured that each GNSS device was positioned in its own pouch specifically designed for the GNSS device. The use of a custom vest has advantages over using a normal vest, where GNSS devices need to be placed on top of one another. Placing GNSS devices on top of one another (Jackson et al., 2018) can obstruct the GNSS antenna, potentially degrading the data quality. Similarly, using a sled for reliability studies (Crang et al., 2021a; Thornton et al., 2019) has its disadvantages, as the GNSS devices are positioned close to the ground and not worn on the human body. This leads to movement patterns that differ from those experienced when the device is worn on the upper back of an athlete, between the shoulders, where they are typically used. The high ICC and low TE values observed in this study suggest that the Catapult Vector S8 GNSS device is reliable, and can confidently be used to compare results of different athletes across training sessions and games to guide informed decision making.

Previous research has shown that different processing practices applied to GNSS data can heavily influence and affect data (Delaney et al., 2019; Delves et al., 2022; Ellens et al., 2024; Thornton et al., 2019; Varley et al., 2017). The results of this study might be superior to comparable previous studies as the processing practises used on the criterion data were the same as those used on the GNSS data. This ensured that any differences present in the data were not due to the processing practices used, but due to differences in the collected raw data.

A limitation of this study was that the data collection was performed in optimal conditions on an open field with unobstructed line of sight to satellites. Although this is applicable to most training sessions for elite team-sports, games usually take place in stadia with high stands and obstructed line of sight to satellites potentially leading to less connected satellites and decreased data accuracy. Future research should conduct similar research in stadia, which may provide less optimal testing conditions. The area of testing was not a full-sized team-based sports pitch, which would have allowed for validation testing during game play. However, to the best of the authors knowledge, there is currently no criterion measure (gold standard) that allows full-sized pitch validation testing.

## Data Availability

Data cannot be shared publicly because of ethical restrictions. Data are available from
the La Trobe Ethics Committee (contact via) for researchers who
meet the criteria for access to confidential data

